# Metabolomics strategy for diagnosing urinary tract infections

**DOI:** 10.1101/2021.04.07.21255028

**Authors:** Daniel B. Gregson, Spencer D. Wildman, Carly C.Y. Chan, Dominique G. Bihan, Ryan A. Groves, Raied Aburashed, Thomas Rydzak, Keir Pittman, Nicolas Van Bavel, Ian A. Lewis

**Author notes:** Corresponding author: Ian A. Lewis Department of Biological Sciences 2500 University Drive NW Calgary, AB, Canada T2N 1N4 1-403-220-4366. Equal contributions (Daniel B. Gregson, Spencer D. Wildman, Carly C.Y. Chan).

## Abstract

Metabolomics has emerged as a mainstream approach for investigating complex metabolic phenotypes but has yet to be integrated into routine clinical diagnostics. Metabolomics-based diagnosis of urinary tract infections (UTIs) is a logical application of this technology since microbial waste products are concentrated in the bladder and thus could be suitable markers of infection. We conducted an untargeted metabolomics screen of clinical specimens from patients with suspected UTIs and identified two metabolites, agmatine and N6-methyladenine, that are predictive of culture positive samples. We developed a 3.2-minute LC-MS assay to quantify these metabolites and showed that agmatine and N6-methyladenine correctly identify UTIs caused by 13 *Enterobacterales* species and 3 non-*Enterobacterales* species, accounting for over 90% of infections (agmatine AUC > 0.95; N6-methyladenine AUC > 0.89). These markers were robust predictors across two blinded cohorts totaling 1,629 patient samples. These findings demonstrate the potential utility of metabolomics in clinical diagnostics for rapidly detecting UTIs.

## Introduction

Infections are caused by a wide range of microbes that, like all organisms, consume nutrients and secrete metabolic waste products. These microbial metabolites can be highly abundant at the site of infection^1^ and thus, could potentially serve as targets for a metabolism-based diagnostic strategy. Urinary tract infections (UTIs) are particularly amenable to this metabolic diagnostic approach because microbial metabolites are concentrated in the bladder and can be easily detected by liquid chromatography mass spectrometry (LC-MS) analysis of patient urine. *Escherichia coli*, the most common UTI pathogen, produces a variety of diamines, polyamines, and acylated conjugates of these molecules that are not typically found in human urine. These molecules have been reported as potential UTI biomarkers for over three decades^2,3^, with more recent work suggesting that these amine-linked molecules may play a role in resistance to nitrosative stress in the bladder^4,5^.

Although microbial metabolites have been recognized as potential diagnostic targets, they have yet to be translated into a clinical diagnostic tool. The reason for this is threefold: 1) concentrations of the microbial polyamines are quite variable in urine^6^ and are thus of borderline utility for diagnostic purposes, 2) only biomarkers of *E. coli* have been reported thus far, which is problematic because it is only one of many UTI-causing organisms^7^, and 3) LC-MS has only recently become sufficiently robust to serve as a practical diagnostic platform. The primary objective of this study was to conduct a systematic analysis of microbial metabolites found in urine from symptomatic UTI patients to identify any molecules with sufficient predictive power to enable rapid, metabolic-based diagnostics. Using an untargeted LC-MS metabolomics approach, we identified two novel biomarkers that report on a wide transect of UTI pathogens. We then assess the potential utility of metabolomics-based UTI screening in two blinded cohorts totaling 1,629 urine samples.

## Results

### UTI biomarker discovery

UTIs are diagnosed based on patient symptoms supported by laboratory diagnostic tests. These clinical tests include culturing urine specimens for 18 hours to identify patients with significant bacteriuria (defined by Alberta Precision Laboratories guidelines as specimens with >10^7^ CFU/L)^8^. Microbial catabolites present in the urine could potentially provide a culture-independent mechanism for rapidly identifying these positive samples. To identify potential biomarkers in these urine specimens, untargeted metabolomics analyses (Figure 1a; Supporting Dataset 1) of 77 culture-positive urine samples and 33 culture negative samples (<10^7^ CFU/L)—all collected from symptomatic patients—were conducted using LC-MS. The most diagnostic features in this dataset, as judged by area under the curve (AUC) of the receiver operating characteristic (ROC) curve, were m/z 114.1025 (AUC=0.98), m/z 131.1289 (AUC=0.89), and m/z 150.0772 (AUC= 0.91; Figure 1b; Supporting Dataset 1). The first two of these untargeted signals were putatively assigned to agmatine, a product of arginine metabolism^9^, and the third was assigned to N6-methyladenine, a modified nucleobase found in prokaryotes^10^. These assignments were validated by LC-MS/MS fragmentation patterns as well as co-elution of the target signal with analytical standards (Figure 1c,d).

**Figure 1.**
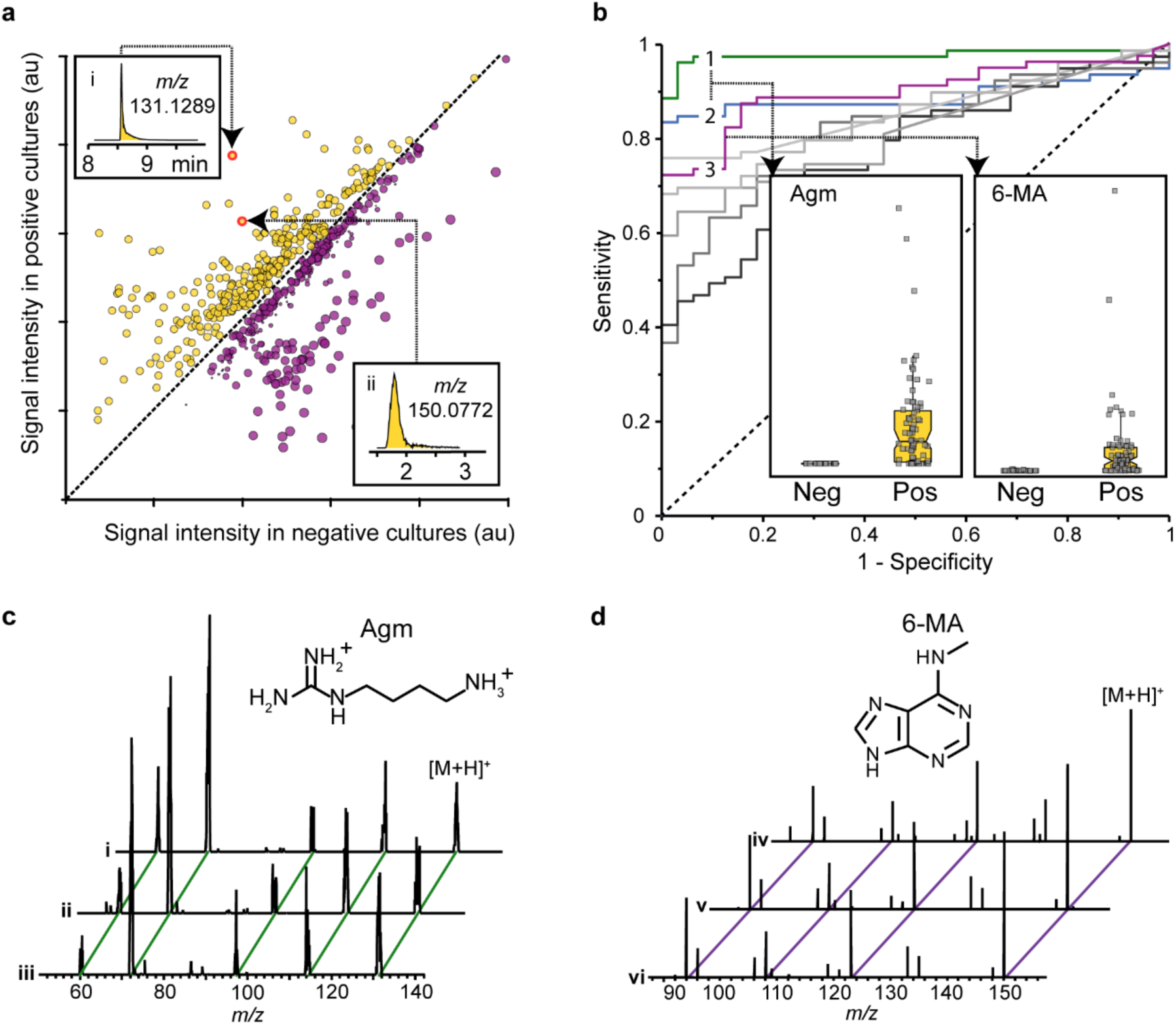
Discovery of agmatine and N6-methyladenine as UTI biomarkers. **(a)** LC-MS data were acquired from metabolites predictive of culture-positive (yellow points) and culture-negative (purple points) urine samples. Potential UTI biomarkers were identified (*m/z* 131.1289 and *m/z* 150.0772 shown as example; see inset for chromatogram). (**b)** Predictive UTI biomarkers were ranked according to receiver operating characteristic (ROC) curves. Signal 1 (*m/z* 114.1025) and signal 2 (*m/z* 131.1289) were assigned to agmatine, and signal 3 (*m/z* 150.0771) was assigned to N6-methyladenine. **(c)** Agmatine and **(d)** N6-methyladenine assignments were verified by tandem LC-MS/MS fragmentation patterns observed in a culture-positive urine sample (i, iv), an analytical standard of the target molecule (ii; 250 nM, v; 50 µM), and a standard added to a culture-negative urine sample at the same concentration (iii, vi). Abbreviations: Agm, agmatine; 6-MA, N6-methyladenine; Neg, growth-negative urine; Pos, growth-positive urine.

### Validation of agmatine as a UTI biomarker

Although both agmatine and N6-methyladenine were predictive of positive urine cultures, the levels of these metabolites were not uniform across UTI-causing pathogens. Infections caused by the most prominent UTI-causing *Enterobacterales* species, such as *E. coli*, were associated with elevated agmatine, whereas N6-methyladenine levels were elevated in infections caused by a small number of non-*Enterobacterales* species (Supporting Dataset 1). To better understand these species-specific phenotypes, the performance of agmatine was assessed using 519 urine cultures that were taken directly from a clinical diagnostic pipeline. Stable isotope labeled [U-^13^C]agmatine was spiked into samples, samples were purified using solid phase extraction (SPE) on a silica column, and agmatine and [U-^13^C]agmatine levels were measured using a targeted mass spectrometry method. Agmatine concentrations were thus quantifiable based on the signal ratio between isotope-labeled and native species. As expected, urinary agmatine levels were closely correlated with the presence of uropathogenic *Enterobacterales*, whereas culture-negative urine samples contained no detectable agmatine (AUC of 0.99, CI of 0.98 - 1.00; Fig 2a; Supporting Dataset 2). Moreover, we found that a diagnostic threshold of 174 nM agmatine was predictive of infections caused by *E. coli*, *Citrobacter* species, *Enterobacter* species*, Klebsiella* species, and *Proteus mirabilis* (sensitivity, 94%; specificity, 97%; PPV, 0.96; NPV, 0.95; Figure 2a), all of which showed significantly higher agmatine levels than urine from culture negative patients (*p* = 2.15×10^-45^, *p* = 1.43×10^-49^, *p* = 2.22×10^-52^, *p* = 3.85×10^-40^, *p* = 5.91×10^-47^, respectively via two-tail unequal variance Welch T-test relative to culture negative urine samples; Supporting Dataset 2).

**Figure 2.**
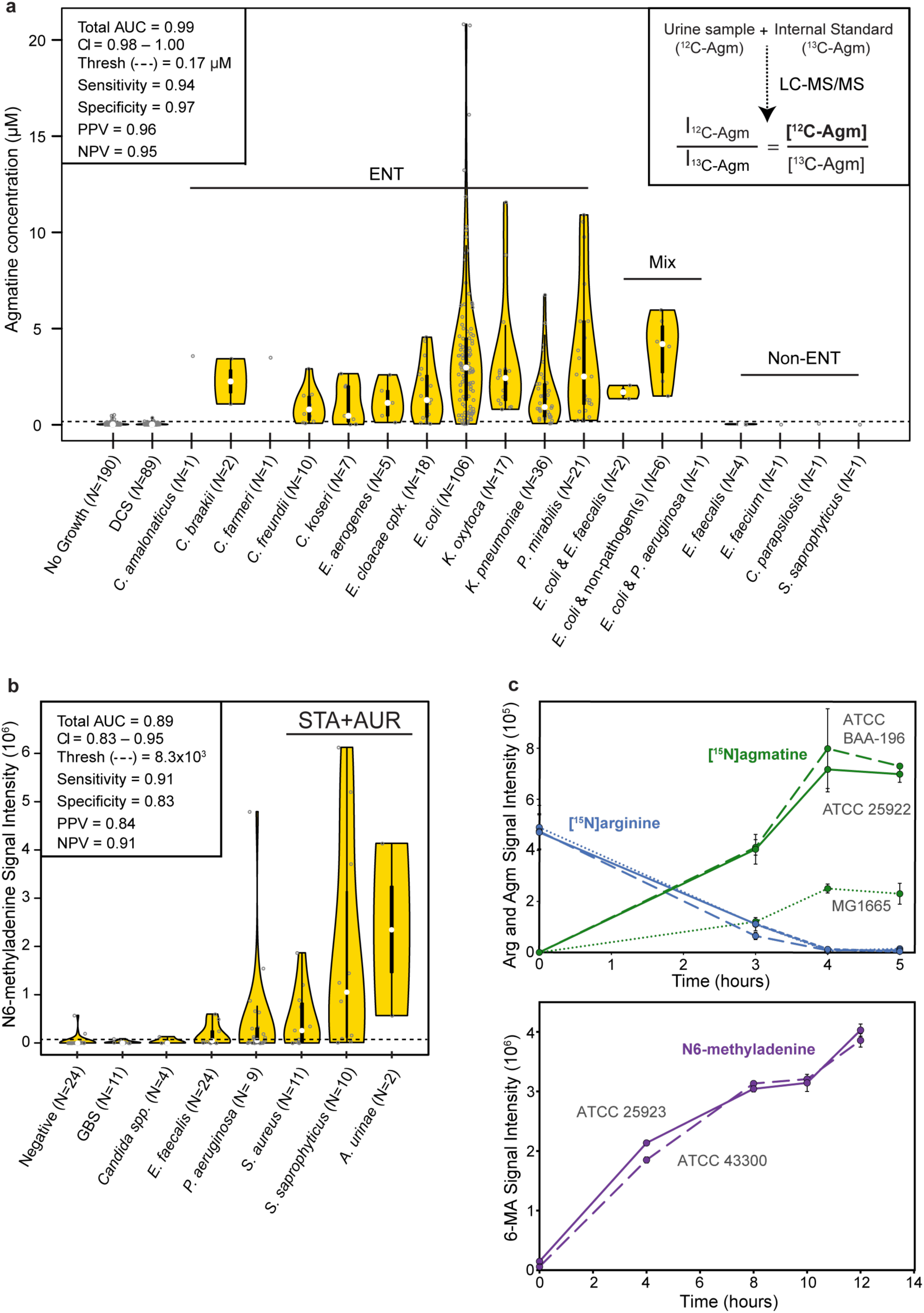
Microbial metabolite signals differentiate *Enterobacterales* and certain non*-Enterobacterales* positive urines from controls. (a) Violin plot of agmatine concentrations in urine samples determined by spike-in of stable isotope-labeled internal standard following SPE (see inset). White dots indicate median values, thick black bars indicate interquartile ranges, and thin black lines indicate 3x interquartile hinge points. **(b)** Violin plot of N6-methyladenine signal intensities from a prospectively collected non-*Enterobacterales* cohort of patient urine samples. **(c)** Three *E. coli* strains grown in sterile urine spiked with [^15^N_2_]arginine for eight hours (top) were monitored for [^15^N_2_]agmatine production. Two *S. aureus* strains grown in sterile urine for 12 hours (bottom) and were monitored for N6-methyladenine production. Abbreviations: Agm, agmatine; AUC, area under curve; CI, confidence interval; Thresh, threshold; PPV, positive predictive value; NPV, negative predictive value; DCS, doubtful clinical significance; Non-ENT, Non-*Enterobacterales*; ENT, *Enterobacterales*; STA, *Staphylococcus* species; AUR, *Aerococcus urinae*; Arg, arginine; 6MA, N6-methyladenine; GBS, Group B Streptococci.

### Validation of N6-methyladenine as a UTI biomarker

Although the agmatine-based screening approach captures most UTI infections caused by *Enterobacterales* species—which collectively account for >83% of UTIs^7^—it did not capture those caused by enterococci, staphylococci, or other less common species (e.g Group B strep, *Pseudomonas aeruginosa*, *Aerococcus urinae*, *Candida albicans*). Since these pathogens account for a small percentage of all UTIs, we prospectively collected a targeted cohort of 71 non-*Enterobacterales* UTI specimens along with 24 negative controls. LC-MS analyses showed that N6-methyladenine signals correctly differentiated culture-negative urine samples from this selected cohort of infections (sensitivity, 91%; specificity, 83%; PPV, 0.84; NPV, 0.91; Figure 2b; Supporting Dataset 3). However, there were clear species-to-species differences in the quality of these metabolic diagnostics, with *Staphylococcus aureus*, *Staphylococcus saprophyticus*, and *Aerococcus urinae* showing the most significant differences after Bonferroni correction from multiple hypothesis testing (*p* = 1.26×10^-3^, *p* = 3.22×10^-4^, *p* = 3.72×10^-6^, respectively via two-tail unequal variance Welch T-test relative to culture negative urine samples; Figure 2b; Supporting Dataset 4). N6-methyladenine levels were predictive of these three non-*Enterobacterales* species (AUC of 0.80; CI, 0.69 – 0.92; Supporting Dataset 3). N6-methyladenine is part of a wider family of adenine derivatives, including 1-methyladenine and adenine, which were elevated in the urine of these UTI patients (Supporting Dataset 4). However, our LC-MS analyses showed that the diagnostic performance of these additional markers was lower, and these compounds were linked to fewer species than N6-methyladenine.

### Validation of microbial metabolic capacity

Agmatine and N6-methyladenine in the urine of UTI patients could potentially originate from microbial or human metabolism. To better understand the origin of these molecules, filter sterilized urine was taken from healthy controls and was inoculated with *E. coli* and *S. aureus* isolates. In the case of the *E. coli* culture, 20 µM of ^15^N-labelled arginine (a physiologically relevant level approximating the concentration of native urinary arginine) was added to the urine at the start of the incubation. Throughout the incubation, isotope-labeled arginine levels progressively decreased while isotope-labeled agmatine was produced (Figure 2c). The appearance of isotope-labeled agmatine is expected under these conditions due to the microbial arginine decarboxylase activity of *E. coli*^9^. Likewise, we screened the *S. aureus* cultures over 12 hours via LC-MS and observed that N6-methyladenine was produced by these microbes when grown in filter-sterilized urine (Figure 2c). In summary, the most diagnostic biomarkers observed in UTI specimens are readily produced by UTI pathogens grown in filter-sterilized urine, indicating that these metabolites are naturally produced through microbial metabolic activity.

Urine specimens for UTI diagnostics are generally collected in outpatient clinics and delivered to centralized microbiology testing labs. Sample transport logistics and other delays in analysis could therefore complicate microbial diagnostics if microbes were allowed to grow. To control this, urine specimens are collected in boric acid tubes, which inhibit microbial growth. To verify that microbes stored in boric acid tubes will not produce our target metabolites, which could cause false positive results in specimens with sub-clinical bacterial levels (< 10^7^ CFU/L), we screened for agmatine production in three reference strains of *E. coli* and four clinical isolates, in boric acid over 24 hours via LC-MS. In the absence of boric acid preservatives, agmatine levels reached a peak intensity after 7 hours and maintained elevated levels over the next 17 hours (Supplementary Figure 1a). In contrast, isolates stored in boric acid preservatives did not produce agmatine over the 24-hour period (Supplementary Figure 1b).

### Clinical Cohort 1

Although untargeted metabolomics is currently not used for clinical diagnostics, mass spectrometry is a routine component of many diagnostic labs where it serves as a platform for quantifying a range of biomolecules including steroid hormones, drugs of abuse, and metabolites that are linked to inborn errors of metabolism^11^. Consequently, much of the requisite infrastructure necessary for diagnosing UTIs via metabolic profiles can be found in diagnostic laboratories. However, the instrumentation used for untargeted metabolomics analyses is quite different from that employed in clinical settings. To demonstrate the feasibility of adapting our metabolomics approach to routine clinical diagnostics, we developed a targeted diagnostics workflow using a triple quadrupole LC-MS instrument that is commonly used for clinical mass spectrometry.

To determine the performance of this clinically-adapted UTI diagnostics approach, we conducted a head-to-head performance evaluation of our LC-MS method versus results obtained via the traditional clinical microbiology approach^12^. This cohort consisted of 587 urine specimens submitted to Alberta Precision Laboratories (South Hub) over a 48-hour period. The demographics of these specimens have been described previously^7^. Briefly, these samples were taken from the greater Calgary metropolitan area where 74% of urine cultures are submitted from ambulatory out-patient visits, 18% from hospitalized patients, and 9% from nursing home residents. Females have a 6-fold higher incidence rate of UTIs than males in this population. Importantly, Alberta follows the “Choosing Wisely Canada” guidelines wherein physicians are instructed to only submit urine cultures from symptomatic patients^13^. This head-to-head trial confirmed our previous results: urine agmatine levels correctly predicted *Enterobacterales* infections (Figure 3, AUC of 0.96; CI of 0.94 – 0.98). Moreover, at the previously determined agmatine threshold of 174 nM, LC-MS diagnostics had a 95% sensitivity and 86% specificity for predicting *Enterobacterales* infections (Figure 3, Supporting Dataset 5). As expected, our culture-negative samples, those classified as being of doubtful clinical significance (DCS; <10^7^ CFU/L), and a small number of urine samples (<5%) containing non-*Enterobacterales* microbes all had agmatine levels below the limit of detection.

**Figure 3.**
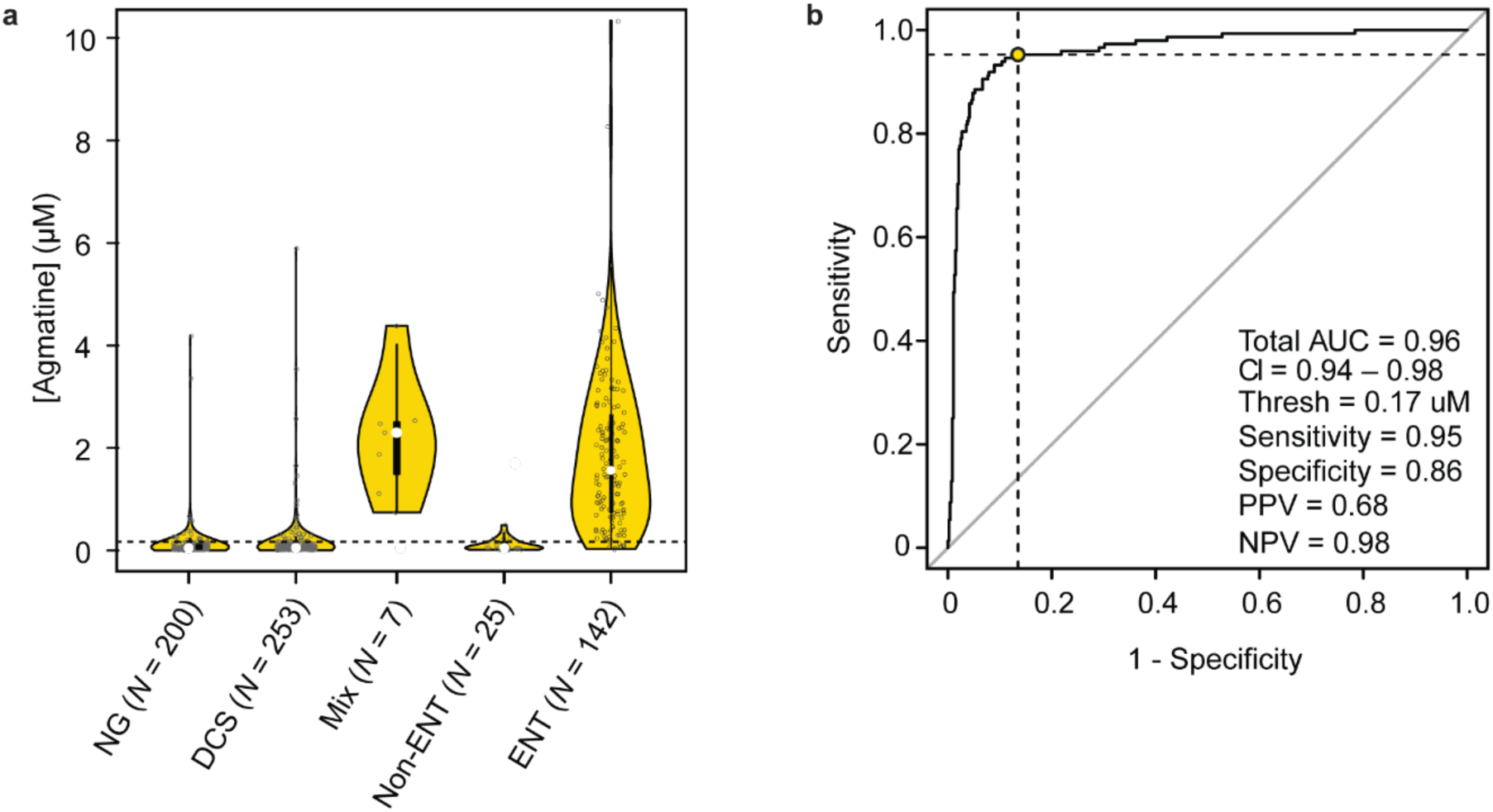
Blinded performance trial of clinically adapted metabolomics-based UTI diagnostics. **(a)** Agmatine concentrations in a blinded cohort of clinical urine samples. Categories on x-axis represent calls from traditional microbiology analysis performed at a regional diagnostic lab. (**b)** Receiver operator curve demonstrating the performance of agmatine as a diagnostic marker for *Enterobacterales* in a blinded, prospective trial. Sensitivity and specificity for 0.17 μM threshold is represented by the yellow dot. Abbreviations: NG, no growth; DCS, doubtful clinical significance; Non-ENT, Non-*Enterobacterales*; ENT, *Enterobacterales*; Mix, polymicrobial cultures containing at least one *Enterobacterales* member; AUC, area under curve; CI, confidence interval; Thresh, threshold; PPV, positive predictive value;NPV, negative predictive value.

### Clinical Cohort 2

One limitation of our original targeted method is that it was specific to agmatine and thus could not be expanded to the analysis of other urinary metabolites, including creatinine, a standard marker of urinary dilution^14,15^. To address this limitation, we developed a “dilute and shoot” sample preparation workflow coupled with a 3.2-minute LC-MS targeted assay for agmatine, N6-methyladenine, and creatinine. To enable accurate quantification of agmatine/N6-methyladenine and creatinine, which are present at substantially different concentrations in urine (nM versus mM), we used a higher dilution (1:200 final) than in the previous trial. Using this assay, we analyzed 1,042 urine specimens from suspect UTI cases. In this cohort, we again showed that urinary agmatine concentrations over 174 nM is an accurate predictor of *Enterobacterales* infections (93% sensitivity; 90% specificity; PPV, 0.67; NPV, 0.98; Figure 4, Supporting Dataset 6). Moreover, we showed that creatinine normalization had a slight negative impact on the performance of the assay (AUC of 0.95, CI 0.93-0.97; versus 0.92, CI 0.89-0.95; for agmatine alone versus creatinine-normalized agmatine levels, respectively; Figure 4, Supplemental Dataset 6). N6-methlyadenine levels were also found to be predictive of UTIs (AUC of 0.71, CI 0.69 – 0.74; inclusive of all species; Supplemental Dataset 6). As expected, the performance of N6-methlyadenine was improved when analyses were restricted to the three known N6-methlyadenine producing species (AUC, 0.89; CI, 0.75 - 1.0; Supporting Dataset 6, Supplementary Figure 2). In this restricted sub-cohort, a N6-methlyadenine concentration of 217 nM had a sensitivity of 0.78 and specificity of 0.79 for differentiating infections with *S. aureus*, *S. saprophyticus*, and *A. urinae* versus infections with all other species and negative specimens (Supplementary Figure 2, Supporting Dataset 6). We also tested the performance of a composite model using both agmatine and N6-methlyadenine levels to predict infections. This combined model performed slightly worse than the agmatine-only model as a generalized tool for detecting UTIs (inclusive of all species, AUC of 0.85 versus 0.78 for the composite versus agmatine-only models, respectively; Supporting Dataset 6).

**Figure 4.**
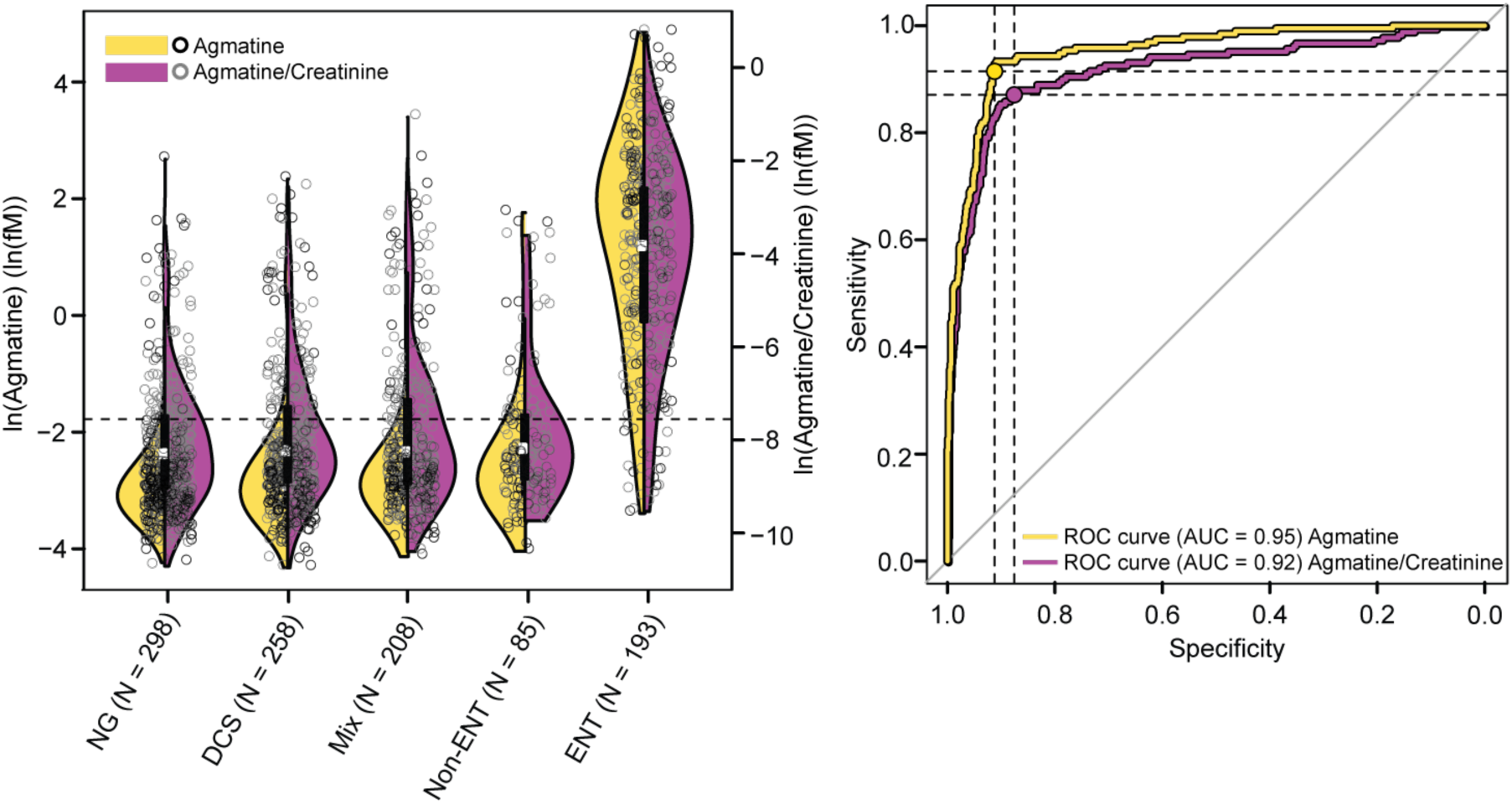
Creatinine normalized agmatine concentrations in a blinded cohort of urine specimens. **(a)** Violin plot of agmatine concentrations compared to creatinine normalized agmatine concentrations. **(b)** Receiver operator curves demonstrating the equivalence between detected agmatine levels and creatinine normalized agmatine levels. Sensitivity and specificity calculated for 0.17 μM threshold. Abbreviations: NG, no growth; DCS, doubtful clinical significance; Non-ENT, Non-*Enterobacterales*; ENT, *Enterobacterales*; Mix, polymicrobial cultures containing at least one *Enterobacterales* member; AUC, area under curve.

## Discussion

UTIs are one of the most common bacterial infections and are responsible for over eight million healthcare visits per year in the U.S. alone^16,17^. As a result, urine cultures are one of the most common tests performed by microbiology reference labs (24–40% of all cultures analyzed)^18^. The current diagnostic workflow used to process these samples follows a multi-day procedure that includes: streaking urines on agar plates, incubation for 18 hours, colony counting, microbial identification via MALDI-TOF mass spectrometry, and antibiotic susceptibility testing^12^. This long diagnostic workflow (24–48 hours) encourages clinicians to prescribe antibiotics before lab results are available. Given that up to 70% of suspected UTI cases collected by clinical testing facilities are ultimately found to be culture negative, the current empiric antimicrobial prescribing practice leads to overprescribing and may contribute to the spread of antimicrobial resistance^19–21^. Therefore, rapid diagnostic tools that can quickly identify patients with UTIs could enable more precise antimicrobial prescribing practices^22–24^.

Herein, we used untargeted metabolomics to identify native biomarkers present in uncultured urine that robustly differentiate between culture negative and culture positive patients. Using these untargeted data, we identified two predictive metabolites (agmatine and N6-methyladenine) that are elevated in uncultured urine from patients who were later confirmed to have UTIs. We show that these biomarkers are produced by UTI pathogens when they are cultured in urine and that an agmatine concentration of 174 nM and N6-methyladenine concentration of 217 nM in urine corresponds with the clinical definition of bacteriuria (>10^7^ CFU/L)^8^. We developed a clinical LC-MS workflow to detect these molecules and demonstrated that these biomarkers are robust predictors of UTIs in two independent cohorts consisting of 1,629 clinical specimens in total (AUC > 0.95 and 0.89 for 13 *Enterobacterales* and 3 non-*Enterobacterales* species, respectively). Collectively, these data show that metabolite levels present in uncultured urine can be used to accurately reproduce the results generated by traditional urine culture and, therefore, could potentially enable novel rapid UTI testing approaches.

The multiple independent trials we conducted over the course of this study provide some insights into leveraging metabolite biomarkers of UTIs. Firstly, our creatinine normalization study showed that correcting metabolite levels to account for variability in urinary dilution does not improve their diagnostic performance – if anything these corrections make things worse (Figure 4; Supporting Dataset 6). This observation is consistent with our microbial culturing experiments, which showed that agmatine and N6-methyladenine are normal microbial catabolites produced when UTI pathogens are grown in urine. Thus, if agmatine and N6-methyladenine levels in patient samples primarily result from microbial metabolism within the urinary tract, we would expect both the biomarker levels and planktonic bacterial density in the urine (in CFU/L) to be diluted in parallel. Given that the clinical definition of bacteriuria is based on bacterial density^8^ it is unsurprising that the microbially-produced catabolites we identified were most diagnostically robust when their levels were not corrected for urinary dilution. This observation has practical implications: quantifying the abundant creatinine necessitated a higher dilution than is necessary for quantifying the low-abundant agmatine and N6-methyladenine. This over-dilution diminished the performance of our target markers in our final clinical cohort, as their signals were closer to the LC-MS noise threshold. Thus, omitting creatinine normalization improves performance of our assay.

Another practical observation from our final clinical cohort is that a mixed agmatine/N6-methyladenine model for predicting infection performed slightly worse than an agmatine only model (all cause UTI; mixed AUC 0.78, 0.74-0.81; agmatine only AUC 0.85, 0.82-0.88; Supporting Dataset 6). The reason for this is that only 9 out of the 1,042 urine specimens (9 out of 277 positive samples; 3.2%) in the final cohort were from infections with N6-methyladenine-producing organisms. This prevalence is consistent with previous studies in the same region (3.2% vs 2.1%)^7^. Thus, screening samples via N6-methyladenine levels added false discovery, but minimal additional sensitivity to the assay. This is a common problem in screening for rare events^25^ and suggests that an agmatine-only screening approach might be preferable.

One important consideration about the generalizability of our findings is that the mix of species observed in clinical specimens will have a direct impact on the clinical performance of our biomarker-based diagnostic approach. The proportions of species we observed are largely consistent with those reported elsewhere with the exception of *S. saprophyticus* (an N6-methyladenine-producer), which was underrepresented relative to the North American average (1% vs 5%)^26,27^. Therefore, N6-methyladenine could potentially be of greater diagnostic value in other health jurisdictions – especially in jurisdictions with a greater proportion of out-patient cases.

The robust performance of agmatine as a diagnostic marker for *Enterobacterales* infections across four separate cohorts suggests that this metabolite is a good candidate for clinical applications. As an LC-MS assay, agmatine screening could be used to rapidly identify patients with clinically significant bacteriuria. This screening approach is 18 hours faster than the traditional urine culture workflow and could accommodate thousands of analyses per day^28^. Alternatively, agmatine screening for UTIs could also be enabled via point-of-care testing tools, such as lateral flow assays, which could be made possible via existing commercial antibodies for agmatine^29^. If such a point-of-care tool were made, it would complement existing dipstick-style UTI screening tests (i.e. nitrate and leucocyte esterase)^30^. In this context, agmatine screening would be both favorable from a performance perspective (Supporting Dataset 7) and because the *Enterobacterales*-specific agmatine test could be used to help guide more precise prescribing decisions.

Although we demonstrate robust performance of our metabolomics-based UTI screening concept, there are some important limitations of this strategy that warrant discussion. Firstly, the biomarkers we identify here report bacteriuria (high levels of bacteria in the urine), not infection. This distinction is important as asymptomatic bacteriuria is common and does not necessarily warrant antimicrobial therapy^31^. Thus, agmatine/N6-methaladenine screening would need to be paired with symptomatic evaluation by a clinician to enable UTI diagnosis (i.e. following existing practice). Secondly, the gold-standard diagnostic workflow involves both species-level identification of the pathogen and empiric antimicrobial susceptibility testing, neither of which is possible with our screening approach alone. However, most UTIs are treated without supporting lab data. Therefore, we envision agmatine/N6-methaladenine screening as a tool to improve empiric prescribing practices rather than as a replacement for culture-based diagnostics. Thirdly, the biomarkers we report here have clinically significant blind spots, especially in the context of UTIs from hospitalised patients. New biomarkers covering *Candida*, *Enterococcus, Pseudomonas,* and group B *Streptococcus* must be identified if our proposed metabolism-based UTI screening concept is to be applied to hospitalized patients. Despite these limitations, the agmatine/N6-methaladenine screening approach we introduce here is robust, reproducible over multiple independent clinical cohorts, and could add significant value as a point-of-care or patient-near tool to enable more precise empiric prescribing decisions for uncomplicated UTIs.

## Methods

### Experimental design

Untargeted analysis for biomarker discovery was performed directly on patient urine samples (77 culture positive, 33 culture negative) using a Q Exactive™ HF Hybrid Quadrupole-Orbitrap™ Mass Spectrometer. Putative biomarkers were identified using MS/MS analysis and matched to standards using fragmentation spectra and retention times. To identify agmatine thresholds that differentiate culture positive (≥10^7^ CFU/L; n=240) versus culture negative urines (<10^7^ CFU/L; n=279), patient urines were spiked with a known concentration of labeled [U-^13^C]agmatine, purified using solid phase extraction (SPE), and analyzed on the TSQ Quantum™ Access MAX Triple Quadrupole Mass Spectrometer. Agmatine concentrations were quantified based on the signal ratio between isotope-labeled and native species. Quantitative analyses of N6-methyladenine were performed directly on patient urine samples (71 culture positive non-*Enterobacterales*, 24 culture negative) using external standards following analysis via the Q Exactive™ HF platform. To demonstrate that agmatine and N6-methyladenine originate from microbial metabolism, *E. coli* and *S. aureus* were incubated in sterilized urine and production of biomarkers was monitored. Similarly, to demonstrate that agmatine concentrations are proportional to microbial load, Mueller Hinton Medium was seeded with varying concentrations of *E. coli* (10^3^-10^8^ CFU/mL) and agmatine levels were measured following a 6-hour incubation period. Inhibition of agmatine production by boric acid was also monitored in filter sterilized urine seeded with *E. coli* during a 24-hour incubation period. Two clinical trials were performed to validate discovered biomarkers. The first clinical trial was performed using SPE purified urine samples (n=587) on the TSQ Quantum™. In the second clinical trial (dilute and shoot approach), methods were developed to quantify agmatine, N6-methyladenine, and creatinine directly in urine using a single injection. For these analyses, urine from residual clinical urine samples (n=1,042) was analyzed after a 1:200 dilution (V:V in 50% methanol final) using the TSQ Altis™ triple quadrupole. The specific LC-MS methods for each experiment are described below.

### Urine sample collection

Mid-stream urine samples were collected from Alberta Precision Laboratories’ (APL) standardized urine analysis workflow, consisting of samples submitted for urine culture from symptomatic ambulatory (74%), hospitalized (18%), and nursing home patients (9%) in the Calgary metropolitan area. Age demographics are divided as follows: 20/1,000 for ages <1; <20/1,000 for ages 1-59; 24/1,000 for ages 60-69; 32/1,000 for ages 70-79; 100/1,000 for ages 80-89; 851/1,000 for ages >90. Incidence rates in females are 30/1,000 and males are 5/1,000. Samples were de-identified prior to transfer to the study team. In brief, urine was plated either manually with a loop or using automated equipment onto UriSelect™ Media (Bio-Rad, Canada) and incubated aerobically for 18 hours. Growth of bacteria was enumerated as per standard recommendations^12^ and colonies present in significant quantities were speciated using the Vitek™ MS system (bioMérieux, Canada). Residual samples from this standard diagnostic lab workflow were fixed 1:1 with 100% methanol within 48 hours post-receipt at APL, and then frozen at -80 °C. All samples were thawed, centrifuged at 14,800 × g, and diluted ten-fold further into 50% HPLC-grade methanol prior to LC-MS analysis. All samples were acquired under approval from Conjoint Health Research Ethics Board certificate REB19-0442.

### Untargeted mass spectrometry

Untargeted analysis of positive and negative urine samples was performed for UTI biomarker discovery and quantification of N6-methyladenine thresholds. Methods were adapted from previously published studies^32–37^. Briefly, metabolic analyses were performed on a Q Exactive™ HF Hybrid Quadrupole-Orbitrap™ Mass Spectrometer (Thermo-Fisher) coupled to a Vanquish™ UHPLC System (Thermo-Fisher). Chromatographic separation of metabolites was performed on Syncronis HILIC UHPLC column (2.1mm x 100 mm x 1.7 µm, Thermo-Fisher) at a flow rate of 600 µL/min using a binary solvent system: solvent A, 20 mM ammonium formate pH 3.0 in mass spectrometry grade H_2_O and solvent B, mass spectrometry grade acetonitrile with 0.1% formic acid (%v/v). For initial biomarker discovery experiments, the following 15-minute gradient was used: 0–2 min, 100% B; 2–7 min, 100–80% B; 7–10 min, 80–5% B; 10–12 min, 5% B; 12–13 min, 5–100% B; 13–15 min, 100% B. The mass spectrometer was run in positive and negative full scan mode at a resolution of 240,000 scanning from 50-750 m/z. Metabolite data were analyzed by El-MAVEN software package^38,39^. Metabolites were identified by matching observed m/z signals (+/- 10 ppm) and chromatographic retention times to those observed from a commercial metabolite library of standards (MSMLS; Sigma-Aldrich). Assignments were verified on the Thermo Fisher Q Exactive™ HF platform via MS/MS fragmentation analysis and co-elution of standards with the identified signals from urine.

### Targeted mass spectrometry

Targeted mass spectrometry analysis for validation of agmatine as a UTI biomarker and clinical trial 1 was performed on a TSQ Quantum™ Access MAX (Thermo Scientific). For quantitative analyses the following transitions were monitored using a fixed collision energy of 15 eV: ^12^C agmatine, 131.2→72.4 m/z; [U-^13^C]agmatine, 136.2→76.4 m/z in positive ionization mode. The same HPLC column and buffers were used as described above, with the following modified 6-minute gradient: 0–0.5 min, 100% B; 0.5–2 min, 100–5% B; 2–3.5 min, 5–0% B; 3.5–4.5 min, 0% B; 4.5–5 min, 0–100% B; 5–6 min, 100% B. For targeted experiments using the TSQ Quantum™, electrospray ionization source conditions were as follows: spray voltage of 3000 V, sheath gas of 25 (arbitrary units), auxiliary gas of 10 (arbitrary units), sweep gas of 0 (arbitrary units), capillary temperature of 275 °C, auxiliary gas temperature of 325 °C.

### Dilute and shoot

For clinical trial 2 (“dilute and shoot” approach), LC-MS/MS analysis was performed on a Vanquish™ UHPLC system, with an integrated Vanquish™ charger module (Thermo Scientific), coupled to a TSQ Altis™ triple quadrupole mass spectrometer (Thermo Scientific) equipped with an electrospray ionization (HESI-II) probe. The UHPLC-MS platform was controlled by an Xcalibur™ data system (Thermo Scientific). Chromatographic separation was achieved on a Syncronis™ HILIC column (50 X 2.1 mm, 1.7 µm, Thermo Fisher Scientific) using a binary solvent system composed of LC-MS grade H_2_O with 20 mM ammonium formate, pH3 (solvent A) and LC-MS grade acetonitrile with 0.1% (%v/v) formic acid (solvent B). The following 3.2 min gradient was used: 0–0.5 min, 95% B; 0.5–1.75 min, 95-30% B; 1.75–1.8 min, 30-5% B; 1.8–2.3 min, 5% B; 2.3-2.4 min, 5-95% B; 2.4-3.2 min, 95% B. The flow rate was 1 mL min^-1^ and the sample injection volume 2 µL. The auto sampler and charger were kept at 6 °C and the column at 30 °C. For quantitative analyses the following transitions were monitored using the following polarities, collision energies and transitions: agmatine, positive, 16 eV, 131.0→97.1 m/z; N6-methyladenine, positive, 23 eV, 150.0→122.9 m/z; creatinine, positive, 13 eV, 114.1→86.1 m/z. Electrospray ionization source conditions were as follows: spray voltage of 3000 V (2500 V, negative mode), vaporizer temperature of 250 °C, sheath gas of 35 (arbitrary units), auxiliary gas flow of 15(arbitrary units) and sweep gas flow of 2 (arbitrary units), capillary temperature of 275°C.

### Determination of agmatine threshold values using [U-^13^C]agmatine

Targeted quantitative analysis was conducted to establish agmatine thresholds between culture positive and culture negative urine samples (Validation of agmatine as a UTI marker) and to assess performance thresholds (Clinical cohort 1). Quantitative analyses of agmatine in 519 urine samples was undertaken using a known concentration of a [U-^13^C]agmatine isotope as an internal standard in the urine sample (see [U-^13^C]agmatine standard biosynthesis and purification below). Prior to MS analysis, samples were purified using a solid phase extraction (SPE) 96-well HyperSep™ Silica plate. Columns were equilibrated with methanol followed by water, then loaded with the sample of interest. Columns were then washed with methanol, water, methanol with 0.1% formic acid, water with 0.1% formic acid. The target analyte was eluted with water containing 2% formic acid. LC-MS analysis was performed using a TSQ Quantum™ Access MAX Triple Quadrupole Mass Spectrometer (Thermo Scientific). For the clinically-adapted blinded evaluation, clinical urine specimens were centrifuged and the supernatant was spiked with the [U-^13^C]agmatine isotope. These samples were analyzed directly on a TSQ Quantum™ Access MAX instrument.

### [U-^13^C]agmatine standard biosynthesis and purification

*Escherichia coli* (strain MG1665) was seeded into M9 minimal media containing 22.2 mM [U-^13^C]glucose, grown overnight, and was then seeded into fresh [U-^13^C]glucose-containing media. This culture was then incubated under agitation in a 37 °C, 5% CO_2_ incubator. Glucose consumption was monitored using a blood glucose monitoring system (Bayer Contour Next). When glucose levels in the media reached 5 mM, the culture was centrifuged, the supernatant was retrieved, was adjusted to pH 7 with ammonium bicarbonate, and was steri-filtered. [U-^13^C]agmatine was isolated and purified using a solid phase extraction column (HyperSep™ Silica Cartridges, # 60108-712, Thermo Scientific), as described above and scaled accordingly to the column volume. The fraction was concentrated to 1/10^th^ of its initial volume in a vacuum concentrator at 4 °C and labeled [U-^13^C]agmatine in the final sample was quantified by stable isotope dilution via LC-MS/MS using an unlabeled internal standard curve.

### Validation of microbial metabolic capacity

*E. coli* and *S. aureus* were inoculated in sterilized urine to verify that agmatine and N6-methyladenine originate from microbial metabolism. *In vitro* microbial samples were cultured from reference strains of bacteria (*E. coli*: MG1665, ATCC 25922, ESBL ATCC BAA-196; *S. aureus*: ATCC25923, ATCC 43300 (MRSA)). Briefly, cryogenic stocks were grown overnight in Mueller-Hinton medium, then inoculated at 10^5^ CFU/mL in sterilized urine and grown at 37 °C. *E. coli* cultures were spiked with [guanido-^15^N_2_]arginine (herein referred to as “[^15^N_2_]arginine”) such that the urine contained an approximate 1:1 ^12^C arginine to [^15^N_2_]arginine ratio. The disappearance of [^15^N_2_]-arginine and appearance of [guanido-^15^N_2_]agmatine (herein referred to as “[^15^N_2_]agmatine”) was monitored over five hours using LC-MS/MS. Similarly, the appearance of the N6-methyladenine signal was assessed in *S. aureus* cultures at 0, 4, 8, 10, and 12 hours with LC-MS.

To demonstrate that agmatine concentrations are proportional to microbial load, Muller Hinton Medium was seeded with varying concentrations of *E. coli* (ATCC 25922; 10^3^-10^8^ CFU/mL) and agmatine levels were measured following a 6-hour incubation period using the Q Exactive™ HF platform. Inhibition of agmatine production by boric acid preservative, at concentrations used in BD Vacutainer Plus Tubes (2.63 mg/mL boric acid, 3.95 mg/mL sodium borate, 1.65 mg/mL sodium formate) was monitored in filter sterilized urine seeded with *E. coli* (three reference strains – MG1665, ATCC 25922, ATCC BAA-196 – and four clinical isolates) over a 24-hour period and analyzed on the Q Exactive™ HF platform.

### Blinded performance evaluation of agmatine versus traditional urine analysis pipeline

Mid-stream urine samples for the blinded head-to-head performance evaluation were collected from Alberta Precision Laboratories’ standardized urine analysis workflow (described above in *Urine Sample Collection*). These samples were stripped of diagnostic information and assigned a unique identifier prior to their transfer to the analytical team. Following analysis using a clinically adapted metabolomics platform, diagnostic data were un-masked and the results of the quantitative agmatine-based MS approach were scored against the clinical diagnostic laboratory calls, using 174 nM as the threshold for UTI-positive samples.

Targeted quantitative assays were conducted in two independently blinded trials using slightly differing methods. In trial 1, targeted mass spectrometry was performed to quantify the levels of agmatine in a blinded cohort of 587 urine specimens. This was done following the targeted mass spectrometry workflow described above using spiked in [U-^13^C]agmatine and SPE sample cleanup. In trial 2, 1,042 urine specimens were analyzed using a simplified dilute and shoot targeted workflow that accurately captures the two target analytes as well as creatinine in a single injection. This approach negates the use of solid phase extraction, and instead, a 1/20 dilution of the sample was directly injected onto the MS following our chromatographic methods described above using a 3.2-minute gradient and external agmatine standard curves were used for quantification.

### Data analysis and statistics

Raw mass spectrometry files were converted into .mzXML files via MSConvertGUI (ProteoWizard Tools)^40^. All full scan MS analyses were conducted in MAVEN. MS/MS data were analyzed using Xcalibur 4.0.27.19 software (Thermo Scientific). Untargeted mass spectrometry data were grouped according to co-variance, co-retention and similarity to common adducts/fragments using previously published software developed in R Statistics^41^. Violin plots, ROC curves, and scatter plots were all generated in R Statistics using existing packages, (FUGU-MS, pROC, and vioPlot). A two-tail unequal variance Welch T-test was used to test differences in agmatine and N6-methyladenine levels in culture positive urine samples compared to negative controls. Alpha thresholds for significance were coerced for multiple hypothesis testing via Bonferroni correction

## Data availability

All data are available in the main text or the supplementary materials. Raw mass spectrometry data are available from the study team upon request.

## Acknowledgments

This work was supported by a Genomics Application Partnership Program award from Genome Canada (10019200), Genome Alberta (10021232), Canadian Institute of Health Research (10020019), and the 2017 Large Scale Applied Research Project competition. This work was made possible in part by a research collaboration agreement with Thermo Fisher Scientific. IAL was supported by an Alberta Innovates Translational Health Chair (10010625). TR was supported by an Eyes High Postdoctoral Fellowship from the University of Calgary (10011121). S.D.W. was supported in part by an NSERC Undergraduate Summer Research Award. Metabolomics data were acquired at the Calgary Metabolomics Research Facility, which is part of the Alberta Centre for Advanced Diagnostics (ACAD; PrairiesCan RIE #22734) and is supported by the International Microbiome Centre and the Canada Foundation for Innovation (CFI-JELF 34986)

## Author Contributions

S.D.W., C.C.Y.C., D.B.G., D.G.B., T.R., and I.A.L. designed and performed the experiments S.D.W., D.B.G., D.G.B., C.C.Y.C., R.A.G., D.B.G., R.A., and T.R. collected and interpreted mass spectrometry data. K.P., N.V.B., and I.A.L. interpreted results wrote the manuscript in consultation with all authors.

