## Supplemental Information for "Metabolomics strategy for diagnosing urinary tract infections"

**Supporting Dataset 1 - Comparative agmatine and N6-methyladenine HPLC-MS signal intensities between UTI-positive and UTI-negative urine in untargeted clinical urine cohort.**

| | | Agmatine | | | N6-Methyladenine | | |
|---|---|---|---|---|---|---|---|
| **Species** | **Number of samples (N)** | **Average Signal Intensity** | **p-value (t-test)*** | **Average Fold Difference*** | **Average Signal Intensity** | **p-value (t-test)*** | **Average Fold Difference*** |
| *E. coli* | 50 | $2.30\times10^{7}$ | $2.48\times10^{-6}$ | 1958 | $1.21\times10^{6}$ | $3.01\times10^{-3}$ | 73.1 |
| *E. faecalis* | 7 | $5.89\times10^{3}$ | 0.0734 | -1.99 | $8.57\times10^{4}$ | $2.77\times10^{-3}$ | 5.18 |
| GBS | 2 | $5.16\times10^{3}$ | 0.258 | -2.27 | $6.52\times10^{4}$ | $7.57\times10^{-3}$ | 3.94 |
| *K. pneumoniae* | 13 | $1.04\times10^{7}$ | $1.96\times10^{-4}$ | 887 | $5.56\times10^{5}$ | $3.84\times10^{-7}$ | 33.6 |
| *P. mirabilis* | 2 | $6.65\times10^{7}$ | $8.56\times10^{-19}$ | 5664 | $8.12\times10^{5}$ | $7.08\times10^{-8}$ | 49.1 |
| *S. aureus* | 3 | $1.65\times10^{4}$ | 0.381 | 1.4 | $1.37\times10^{6}$ | $2.11\times10^{-9}$ | 83 |
| All Negatives | 33 | $1.17\times10^{4}$ | N/A | N/A | $1.65\times10^{4}$ | N/A | N/A |

**compared to negative*

### Supporting Dataset 2 - Agmatine concentrations in prospective UTI-positive and UTI-negative urine samples collected from clinical diagnostic pipeline

| Species | | Agmatine Concentration (μM) | p-value (t-test)* | Average Fold Change* |
|---|---|---|---|---|
| ***Citrobacter* species** | **21** | 1.37 | $1.43\times10^{-49}$ | 31.59 |
| *C. amalonaticus* | 1 | 3.57 | N/A | 82.39 |
| *C. braakii* | 2 | 2.26 | $4.40\times10^{-78}$ | 52.02 |
| *C. farmeri* | 1 | 3.46 | N/A | 80.44 |
| *C. freundii* | 10 | 0.97 | $1.88\times10^{-45}$ | 22.44 |
| *C. koseri* | 7 | 1.07 | $2.65\times10^{-39}$ | 24.59 |
| ***Enterobacter* species** | **23** | 1.56 | $2.22\times10^{-52}$ | 36.04 |
| *E. aerogenes* | 5 | 1.22 | $6.55\times10^{-54}$ | 28.13 |
| *E. cloacae* | 18 | 1.66 | $5.63\times10^{-52}$ | 38.24 |
| ***E. coli*** | **106** | 3.72 | $2.15\times10^{-45}$ | 85.82 |
| ***Klebsiella* species** | **53** | 2.03 | $3.85\times10^{-40}$ | 46.75 |
| *K. oxytoca* | 17 | 3.08 | $3.72\times10^{-48}$ | 71.06 |
| *K. pneumoniae* | 36 | 1.53 | $9.64\times10^{-44}$ | 35.27 |
| ***P. mirabilis*** | **21** | 3.57 | $5.91\times10^{-47}$ | 82.23 |
| ***E. coli & E. faecalis*** | **2** | 1.7 | $1.23\times10^{-99}$ | 39.18 |
| ***E. coli & P. aeruginosa*** | **1** | 1.03 | N/A | 23.83 |
| ***E. coli & non-pathogenic*** | **6** | 3.92 | $2.79\times10^{-116}$ | 90.33 |
| ***C. parapsilosis*** | **1** | 0.06 | N/A | 1.28 |
| ***E. faecalis*** | **4** | 0.03 | 0.019 | -1.34 |
| ***E. faecium*** | **1** | 0.01 | N/A | -4.33 |
| ***S. saprophyticus*** | **1** | 0.01 | N/A | -5.04 |
| **All Negatives (including DCS)** | **279** | 0.04 | N/A | N/A |

**compared to negative*

**Supporting Dataset 3 – Performance indicators of N6-methyladenine for all non-*Enterobacterales* species and the N6-methyladenine producing species in prospective UTI-positive and UTI-negative urine samples.**

| Biomarker | Target Species | N1 (target sp.) | N2 | Threshold (Signal intensity) | AUC | CI | Sensitivity | Specificity | PPV | NPV |
|---|---|---|---|---|---|---|---|---|---|---|
| N6-methyladenine | Non-*Enterobacterales* species | 71 | 24 | $8.13 \times 10^3$ | 0.89 | 0.83 – 0.95 | 0.91 | 0.83 | 0.84 | 0.91 |
| N6-methyladenine | *S. saprophyticus*, *S. aureus*, *A. urinae* | 23 | 72 | $7.1 \times 10^4$ | 0.80 | 0.69 – 0.92 | 0.74 | 0.75 | 0.49 | 0.90 |

### Supporting Dataset 4 - Agmatine and N6-methyladenine HPLC-MS signal intensities between UTI-positive and UTI-negative urine samples.

| | | Agmatine | | | N6-Methyladenine | | | | 1-Methyladenine | | | Adenine | | |
|---|---|---|---|---|---|---|---|---|---|---|---|---|---|---|
| Species | Number of urine samples (N) | Average Signal Intensity | p-value (t-test)* | Average Fold Difference * | Average Signal Intensity | Confidence Interval | p-value (t-test)* | Average Fold Difference* | Average Signal Intensity | p-value (t-test)* | Average Fold Difference* | Average Signal Intensity | p-value (t-test)* | Average Fold Difference* |
| *A. urinae*† | 2 | $1.81\times10^{3}$ | 0.616 | -2.37 | $2.33\times10^{6}$ | $-1.14\times10^{6} – 5.85\times10^{6}$ | $3.72\times10^{-6}$ | 62.88 | $1.41\times10^{8}$ | $1.33\times10^{-25}$ | 126.57 | $2.38\times10^{7}$ | 0.000324 | 8.04 |
| *C. albicans* | 3 | 0 | 0.29 | N/A | $4.24\times10^{4}$ | $-3.58\times10^{4} – 1.29\times10^{5}$ | 0.942 | 1.14 | $1.39\times10^{6}$ | 0.759 | 1.25 | $3.58\times10^{6}$ | 0.864 | 1.21 |
| *C. glabrata* | 1 | $7.88\times10^{3}$ | N/A | 1.84 | 0 | N/A | N/A | N/A | 0 | N/A | N/A | $1.77\times10^{7}$ | N/A | N/A |
| *E. faecalis* | 24 | $5.53\times10^{5}$ | 0.317 | 129 | $4.10\times10^{5}$ | $9.32\times10^{3} – 8.13\times10^{5}$ | 0.0769 | 11.06 | $1.82\times10^{7}$ | 0.143 | 16.33 | $9.62\times10^{6}$ | 0.0482 | 3.25 |
| GBS | 11 | $7.08\times10^{3}$ | 0.352 | 1.65 | $2.28\times10^{4}$ | $3.82\times10^{3} – 3.77\times10^{4}$ | 0.702 | -1.63 | $1.57\times10^{6}$ | 0.502 | 1.41 | $3.79\times10^{6}$ | 0.68 | 1.28 |
| *P. aeruginosa* | 9 | $3.41\times10^{4}$ | 0.155 | 7.94 | $1.54\times10^{5}$ | $1.41\times10^{4} – 3.15\times10^{5}$ | 0.0531 | 4.15 | $3.08\times10^{7}$ | 0.063 | 27.56 | $1.32\times10^{7}$ | 0.0494 | 4.45 |
| *S. aureus*† | 11 | $2.38\times10^{4}$ | 0.165 | 5.55 | $4.92\times10^{5}$ | $1.32\times10^{5} – 8.60\times10^{5}$ | $1.26\times10^{-3}$ | 13.25 | $4.60\times10^{7}$ | 0.014 | 41.25 | $1.47\times10^{7}$ | 0.00715 | 4.97 |
| *S. saprophyticus*† | 10 | $2.11\times10^{3}$ | 0.33 | -2.03 | $1.87\times10^{6}$ | $4.72\times10^{5} – 3.31\times10^{6}$ | $3.22\times10^{-4}$ | 50.5 | $1.65\times10^{7}$ | 0.002 | 14.77 | $5.17\times10^{6}$ | 0.293 | 1.74 |
| All Negatives | 24 | $4.29\times10^{3}$ | N/A | N/A | $3.71\times10^{4}$ | $-1.24\times10^{4} – 8.43\times10^{4}$ | N/A | N/A | $1.12\times10^{6}$ | N/A | N/A | $2.96\times10^{6}$ | N/A | N/A |

**compared to negative*

† *Species found to be significant relative to negative growth after Bonferroni correction*

**Supporting Dataset 5 - Agmatine concentrations in blinded prospective performance trial.**

| | | Agmatine | | |
|---|---|---|---|---|
| Species | Number of urine samples (N) | Average Concentration (µM) | p-value (t-test)* | Average Fold Change* |
| ***Enterobacterales* species** | 98 | 1.92 | $2.76\times10^{-69}$ | 13.39 |
| *C. freundii* | 2 | 1.26 | $5.26\times10^{-4}$ | 8.74 |
| *E. aerogenes* | 2 | 1.11 | $2.26\times10^{-3}$ | 7.73 |
| *E. cloacae* | 5 | 0.73 | $3.90\times10^{-3}$ | 5.08 |
| *E. coli* | 73 | 2.02 | $7.30\times10^{-77}$ | 14.03 |
| *K. oxytoca* | 3 | 0.9 | $3.30\times10^{-3}$ | 6.29 |
| *K. pneumoniae* | 8 | 1.61 | $9.53\times10^{-18}$ | 11.23 |
| *P. mirabilis* | 4 | 4.17 | $3.09\times10^{-39}$ | 29.02 |
| *P. rettgeri* | 1 | 0.76 | N/A | 5.29 |
| *C. freundii & E. faecalis* | 1 | 1.11 | N/A | 7.75 |
| *E. coli & E. faecalis* | 3 | 3.25 | $2.13\times10^{-28}$ | 22.65 |
| *E. coli & K. pneumoniae* | 1 | 1.88 | N/A | 13.06 |
| *E. coli & P. mirabilis* | 2 | 2.56 | $5.60\times10^{-13}$ | 17.84 |
| *E. coli & S. viridans* | 1 | 2.47 | N/A | 17.22 |
| **non-*Enterobacterales* species** | 25 | 0.11 | 0.682 | -1.34 |
| *A. urinae* | 1 | 0.04 | N/A | -3.85 |
| *C. albicans* | 2 | 0.13 | 0.966 | -1.1 |
| *E. faecalis* | 14 | 0.08 | 0.574 | -1.87 |
| *E. faecium* | 1 | 0.04 | N/A | -3.29 |
| GBS | 2 | 0.04 | 0.747 | -3.4 |
| *P. aeruginosa* | 3 | 0.15 | 0.972 | 1.06 |
| *E. faecalis & C. albicans* | 1 | 0.22 | N/A | 1.56 |
| *E. faecalis & S. aureus* | 1 | 0.49 | N/A | 3.44 |
| **All Negatives (including DCS)** | 456 | 0.14 | N/A | N/A |

** compared to negative*

**Supporting Dataset 6 – Performance of agmatine and N6-methyladenine in the second clinical trial for predicting bacteriuria (n = 1,042)†**

| Biomarker | Target Species | N1 (Target sp.) | N2 | OptThresh (nM) | AUC | CI | Sensitivity | Specificity | PPV | NPV |
|---|---|---|---|---|---|---|---|---|---|---|
| Agmatine | *Enterobacterales* species | 192 | 846 | 174[1] | N/A | N/A | 0.93 | 0.90 | 0.67 | 0.98 |
| Agmatine | *Enterobacterales* species | 192 | 846 | 202 | 0.95 | 0.93 - 0.97 | 0.91 | 0.91 | 0.71 | 0.98 |
| Agmatine | *Enterobacterales* and Non-*Enterobacterales* species | 277 | 761 | 85.0 | 0.85 | 0.82 - 0.88 | 0.77 | 0.78 | 0.56 | 0.90 |
| Agmatine: Creatinine | *Enterobacterales* species | 192 | 846 | 0.0013‡ | 0.92 | 0.89 - 0.95 | 0.87 | 0.88 | 0.62 | 0.97 |
| N6-methyladenine | *Enterobacterales* species | 192 | 846 | 139 | 0.73 | 0.69 - 0.77 | 0.68 | 0.68 | 0.33 | 0.90 |
| N6-methyladenine | Non-*Enterobacterales* species | 85 | 953 | 139 | 0.58 | 0.51 - 0.64 | 0.56 | 0.58 | 0.11 | 0.94 |
| N6-methyladenine | *Enterobacterales* and Non-*Enterobacterales* species | 277 | 761 | 139 | 0.71 | 0.69 - 0.74 | 0.66 | 0.66 | 0.41 | 0.84 |
| N6-methyladenine | *S. saprophyticus*, *S. aureus*, *A. urinae* | 9 | 1029 | 217 | 0.89 | 0.75 - 1.00 | 0.78 | 0.79 | 0.03 | 1.00 |
| Agmatine:N6-methyladenine | *Enterobacterales* and Non-*Enterobacterales* species | 277 | 761 | 174 : 217 | 0.78 | 0.74 - 0.81 | 0.77 | 0.78 | 0.57 | 0.90 |

**†** Agmatine (Agm), N6-methyladenine (6MA), and creatinine (Cre) concentrations in the urine of suspect UTI cases were quantified by LC-MS. Agm, 6MA, or the Agm/Cre ratios were then used to predict culture positive versus culture negative samples (as judged relative to gold-standard urine culture methods) based on pre-determined thresholds. Optimal thresholds (OptThresh), as judged based on the area under the receiver operator characteristic (AUC), were defined as the point that maximized both sensitivity and specificity. Standard error (SE) and confidence interval (CI) were calculated for all AUC values. Sensitivity, specificity, positive predictive value (PPV), negative predictive value (NPV) report performance figures at the predetermined thresholds. Target species describes the microbial taxa that were defined as true positives.

[1] Predetermined threshold chosen differed from the optimal threshold provided by the AUC.

‡ Unitless, reports a ratio between agmatine and creatinine

**Supporting Dataset 7 – Performance of agmatine screening relative to dipstick UTI screening tools. Adapted from Semeniuk and Church (J. Clin. Microbiol. 1999, P. 3051-3052)**

| | Sensitivity (%) at CFU/mL: | | Specificity (%) at CFU/mL: | | Predictive Value (%) at colony counts of: | | | |
|---|---|---|---|---|---|---|---|---|
| | | | | | PPV | | NPV | |
| | $10^4$ | $10^5$ | $10^4$ | $10^5$ | $10^4$ | $10^5$ | $10^4$ | $10^5$ |
| LE | 76.9 | 84.4 | 59.4 | 59.4 | 16.0 | 19.4 | 96.3 | 97.1 |
| NIT | 7.9 | 43.6 | 96.6 | 96.6 | 27.3 | 75.0 | 86.3 | 88.2 |
| LE and NIT | 26.0 | 84.0 | 98.3 | 98.3 | 42.9 | 84.0 | 96.3 | 98.3 |
| Agmatine screen | 86.7 | 96.6 | 99.5 | 99.5 | 22.0 | 78.9 | 99.5 | 99.2 |
| Agmatine screen (Ent only) | 96.2 | 99.1 | 99.7 | 99.7 | 36.8 | 89.6 | 99.7 | 99.7 |

Abbreviations: LE, leukocyte esterase; NIT, nitrite; Ent, *Enterobacterales*; PPV, positive predictive value; NPV, negative predictive value

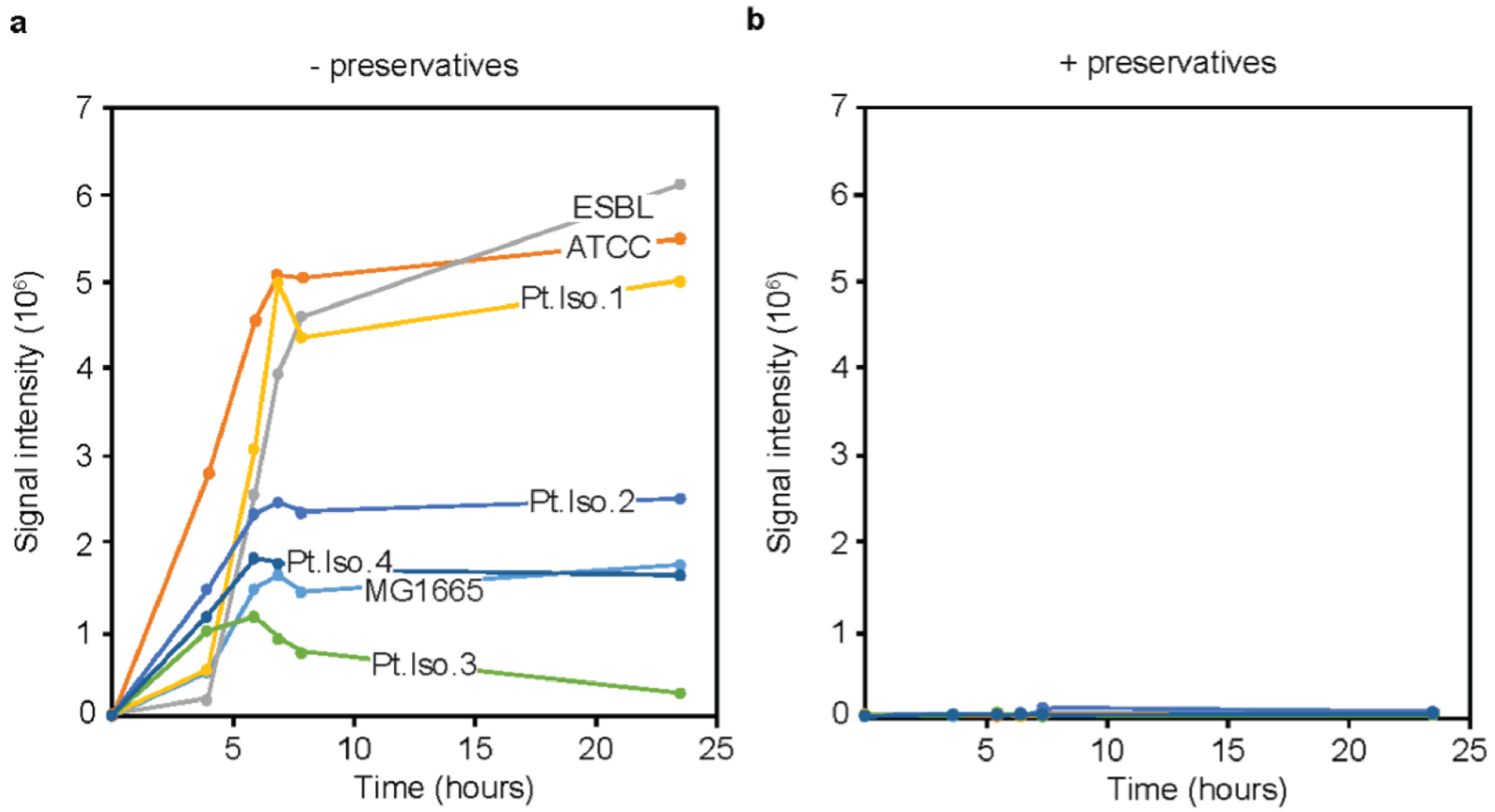


**Figure S1. Boric acid preservatives eliminate agmatine production. (a)** Seven strains of E.coli (MG1665, ATCC 25922, ESBL ATCC BAA-196, and 4 clinical isolates; Pt. Iso 1–4) were subcultured in filter-sterilized urine without or **(b)**, with the preservative cocktail found in conventional clinical urine sample collection tubes. The agmatine produced in these cultures was quantified by LC-MS.

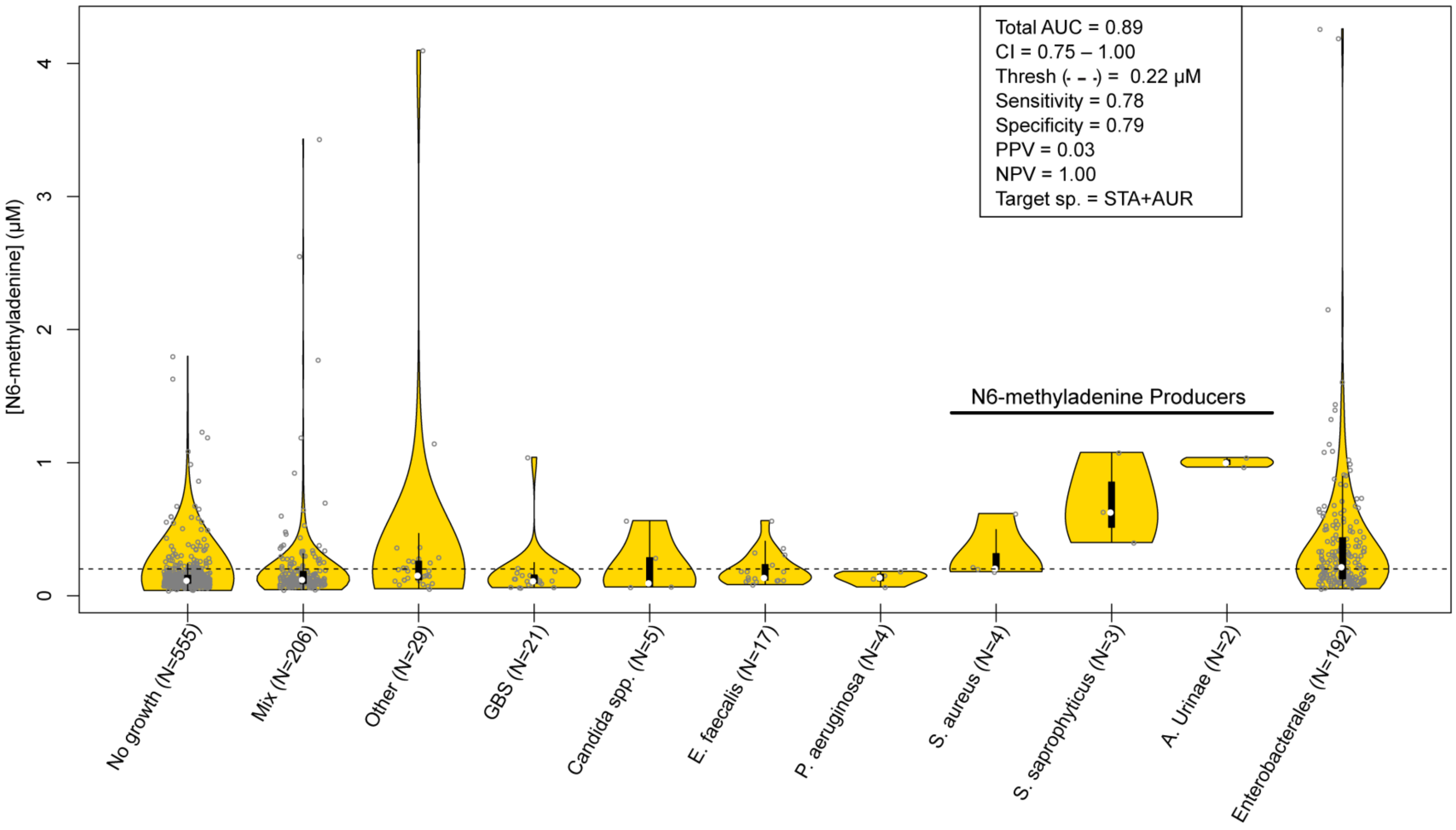


**Figure S2. Blinded performance trial of N6-methyladenine with clinical UTI specimens.** Violin plot of N6-methyladenine concentrations in a blinded cohort of 1,038 clinical urine samples**.** Sensitivity, specificity, PPV, and NPV calculated for the N6-methyladenine producing species at a 0.22 μM threshold. Abbreviations: STA, *Staphylococcus* species; AUR, *Aerococcus urinae*; AUC, area under curve; CI, confidence interval; Thresh, threshold; PPV, positive predictive value; NPV, negative predictive value.
